## Supplementary Figure for "Estimating the long-term health impact of nicotine exposure by dissecting the effects of nicotine versus non-nicotine constituents of tobacco smoke: A multivariable Mendelian randomisation study"

**Supplementary Figures**

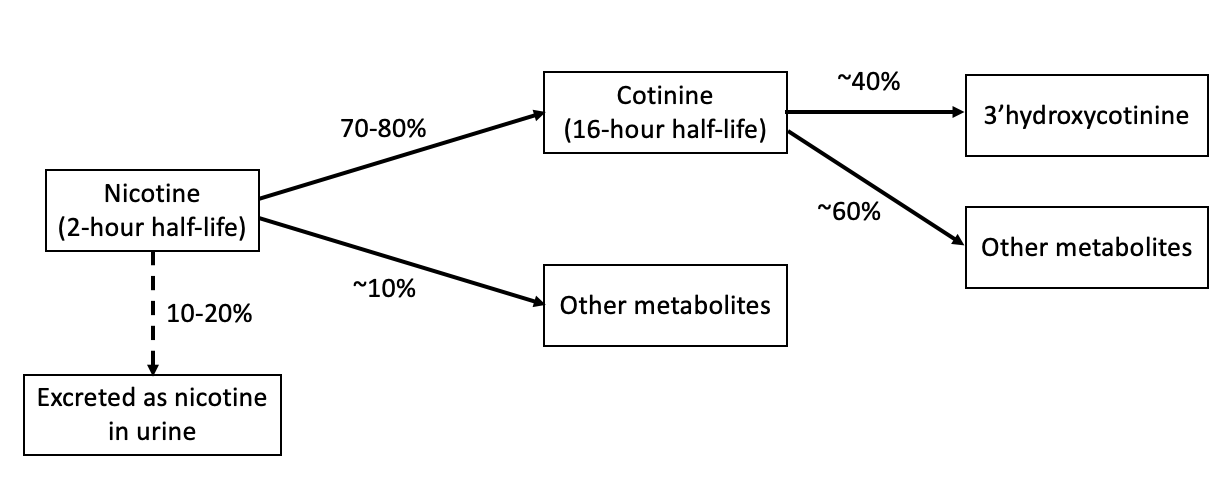

Supplementary Figure S1. A diagram to show the by-products of nicotine metabolism. Adapted from (Benowitz, Hukkanen, & Jacob, 2009)

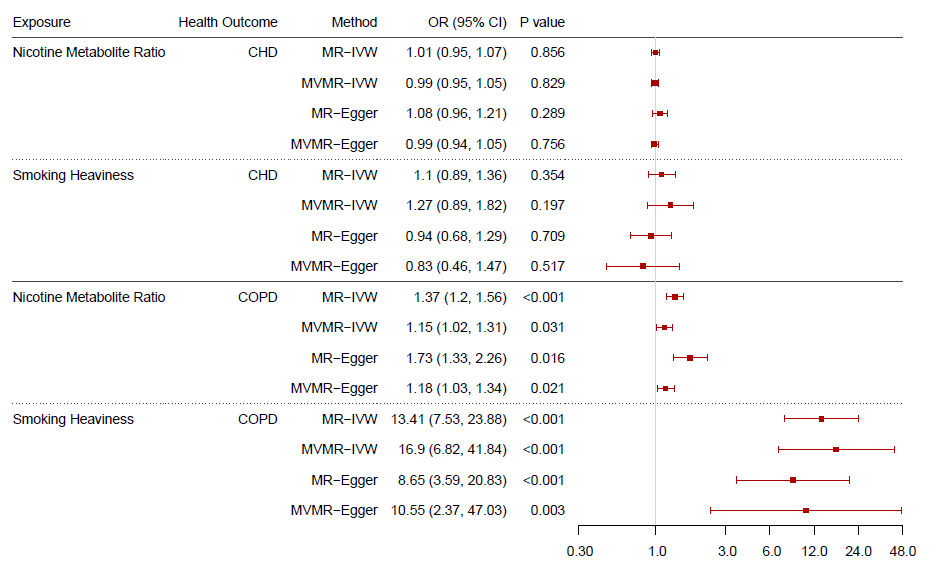

Supplementary Figure S2. Forest plot displaying the effect of nicotine metabolite ratio and smoking heaviness on coronary heart disease (CHD) and chronic obstructive pulmonary disease (COPD): univariable Mendelian randomisation (MR) and multivariable Mendelian randomisation (MVMR) results among current smokers.

**References**

Benowitz, N. L., Hukkanen, J., & Jacob, P., 3rd. (2009). Nicotine chemistry, metabolism, kinetics and biomarkers. *Handb Exp Pharmacol*(192), 29-60. doi:10.1007/978-3-540-69248-5_2
