## Supplemental Note for "Estimating the long-term health impact of nicotine exposure by dissecting the effects of nicotine versus non-nicotine constituents of tobacco smoke: A multivariable Mendelian randomisation study"

**Supplementary Material**

**Supplemental Note S1.**

Buchwald and colleagues (2020) reported summary-level statistics from a genome-wide association study (GWAS) meta-analysis of cotinine plus 3’hydroxycotinine (COT+3HC). Single nucleotide polymorphisms (SNPs) were reported as independent if they explained additional variance in a step-wise conditional regression using genome-wide complex trait analysis (Yang, Lee, Goddard, & Visscher, 2011). The reported data is available on request from the authors. Ware et al. (2016) report summary-level statistics from a GWAS meta‑analysis of cotinine levels (per standard deviation change) among 4,548 daily smokers of European Ancestry (data available at: <https://doi.org/10.5523/bris.182rhz19hg3lz1172a7yfcap9v>). SNPs were reported as independent if they reached genome-wide significance using an iterative process of conditional analyses.

**Supplemental Note S2.**

In UK Biobank, 49,979 individuals were genotyped using the UK BiLEVE array and 438,398 using the UK Biobank axiom array. Pre-imputation QC, phasing and imputation are described elsewhere (Bycroft et al., 2018). In brief, prior to phasing, multiallelic SNPs or those with minor allele frequency (MAF) ≤1% were removed. Phasing of genotype data was performed using a modified version of the SHAPEIT2 algorithm (O'Connell et al., 2016). Genotype imputation to a reference set combining the UK10K haplotype and HRC reference panels (Huang et al., 2015) was performed using IMPUTE2 algorithms (Howie, Marchini, & Stephens, 2011). The analyses presented here were restricted to autosomal variants within the HRC site list using a graded filtering with varying imputation quality for different allele frequency ranges. Therefore, rarer genetic variants are required to have a higher imputation INFO score (Info>0.3 for MAF >3%; Info>0.6 for MAF 1-3%; Info>0.8 for MAF 0.5-1%; Info>0.9 for MAF 0.1-0.5%) with MAF and Info scores having been recalculated on an in-house derived ‘European’ subset (Mitchell et al., 2018).

Individuals with sex-mismatch (derived by comparing genetic sex and reported sex) or individuals with sex-chromosome aneuploidy were excluded from the analysis (n=814). We restricted the sample to individuals of ‘European’ ancestry as defined by an in-house k-means cluster analysis performed using the first 4 principal components provided by UK Biobank using R software environment. The largest cluster from the in-house k-means cluster analysis was n=464,708. To model population structure in the sample, we used 143,006 directly genotyped SNPs, obtained after filtering on MAF > 0.01; genotyping rate > 0.015; Hardy-Weinberg equilibrium p-value < 0.0001 and LD pruning to an r2 threshold of 0.1 using PLINKv2.00.

**Supplemental Note S3.**

We identified Chronic Obstructive Pulmonary Disease (COPD) cases as participants who responded that they had been diagnosed with COPD in response to the question "Has a doctor ever told you that you have had any of the conditions below?".

Forced expiratory volume in 1 second (FEV-1) and forced vital capacity (FVC) were measured multiple times using a Vitalograph spirometer. We used the ‘best measure’ of both FEV-1 and FVC which were identified as the highest value recorded with no measurement issues reported.

Coronary heart disease (CHD) diagnosis was determined using linked hospital admission data with ICD codes relating to Ischemic Heart Disease (ICD-9 410-414; ICD-9 4100-4149; ICD-10 I20-I25). The measure included angina pectoris, acute myocardial infarction, subsequent myocardial infarction, certain current complications following acute myocardial infarction, other acute ischaemic heart diseases, and chronic ischaemic heart disease.

Heart rate (beats per minute) was assessed on multiple occasions per session. Heart rate can be affected by numerous factors such as exercise (Evans, 1985) and stress (Kim, Cheon, Bai, Lee, & Koo, 2018). To allow time for the participant’s heart rate to normalise during the session, we used the second measure taken within the session.

***UK Biobank health outcome field codes***

Field IDs can be entered into the online variable search platform where information can be found on measurement (available at <http://biobank.ndph.ox.ac.uk/showcase/search.cgi>).

Chronic Obstructive Pulmonary Disease: 22130

Forced Expiratory Volume and Forced Vital Capacity: 20150 and 20151

Coronary Heart Disease: 41270 and 41271

Heart rate: 102

**Supplemental Note S4.**

While harmonising the exposure datasets with the lung cancer outcome dataset, two SNPs associated with cigarettes per day (CPD) were removed due to having intermediate allele frequencies (rs1737894, rs28438420). Two SNPs were associated with both NMR and CPD (rs56113850, rs117824460), and one SNP associated with NMR was not available in the CPD GWAS dataset (rs34638591). The p-value for the rs117090198 SNP-exposure effect size was unusually high prior to the conditional analysis in the original GWAS (Buchwald et al., 2020) and the original authors could not provide a definitive explanation for this. We removed this SNP to reduce heterogeneity. Where SNPs associated with the nicotine metabolite ratio (NMR) or CPD were not available in either of the two other datasets, we searched for proxy SNPs with a minimum linkage disequilibrium (LD) R^2^ of 0.8. In MVMR analyses, all SNPs included in the model should be independent of each other (i.e., the SNPs associated with NMR must also be independent of the SNPs associated with CPD and vice versa). To ensure overall independence, we clumped the full list of SNPs (N = 59 SNPs, LD R^2^ < 0.1, clumping window > 500 kb). Given the limited number of SNPs associated with the NMR, SNPs associated with CPD were dropped from the analysis rather than SNPs associated with NMR to preserve instrument strength. Clumping removed 20 SNPs.

**Supplemental Note S5.**

We used four complimentary univariable Mendelian randomisation (MR) methods (inverse variance weighted [IVW], MR-Egger, weighted median-based estimation and weighted modal-based estimation) (Bowden, Davey Smith, & Burgess, 2015; Bowden, Davey Smith, Haycock, & Burgess, 2016; Burgess, Butterworth, & Thompson, 2013; Hartwig, Davey Smith, & Bowden, 2017). Using a variety of Mendelian randomisation (MR) methods with different assumptions with respect to horizontal pleiotropy – which occurs when single genetic variants influence multiple phenotypes (Davey Smith & Hemani, 2014) – allows us to better understand whether the effect of our exposure on our outcome is causal. Consistent results across these methods provide stronger evidence to support a true causal effect which is not the result of a false positive (Lawlor, Tilling, & Davey Smith, 2016). We also estimated the weighted regression dilution (I^2^_GX_) for each MR-Egger analysis (Bowden, Del Greco, et al., 2016) and applied simulation extrapolation SIMEX (Lederer & Küchenhoff, 2006) corrections to MR-Egger analysis where I^2^_GX_ estimates were below 0.9 (which would indicate the effect estimate is biased by 10% due to measurement error) (Bowden, Del Greco, et al., 2016). This analysis was first restricted to ever smokers to capture the long-term effects of smoking/nicotine use (i.e., including former smokers who may have quit smoking due to developing a health issue), and then further restricted to current smokers only (where the data were available) to explore the potential effects of smoking cessation (i.e., recoverable effects). If a poor health outcome was found among current smokers but not ever smokers, it would indicate that health outcome may improve following smoking cessation. Restricting the analysis to former smokers, we can further explore this potential effect. Finally, the analysis was restricted to never smokers to explore the potential presence of pleiotropic pathways and bias due to population stratification.

**Supplemental Note S6.**

There are three main assumptions which underlie instrumental variable (IV) analyses which are applicable to MR analyse:

1. Relevance – the genetic variant(s) used as IVs must be associated with the exposure of interest.
2. Independence – the genetic variant(s) must not share any unmeasured cause with the outcome (e.g., via population stratification, collider bias, dynastic effects, or assortative mating).
3. Exclusion restriction – the genetic variant(s) must not affect the outcome except through its potential effect on the exposure of interest.

**Supplemental Note S7.**

These results are also shown in Supplementary Figure S2, Supplementary Table S9, and Supplementary Table S10.

***CHD***

Among current smokers, there is no clear evidence to suggest that either NMR or smoking heaviness affect CHD risk (OR = 1.01, 95% CI 0.95 to 1.07, OR = 1.10 95% CI 0.89 to 1.36 respectively), nor is there evidence to suggest a clear effect of NMR or smoking heaviness in the IVW-MVMR analysis (OR = 0.99, 95% CI 0.95 to 1.05, OR = 1.27 95% CI 0.89 to 1.82 respectively). There is considerable evidence of heterogeneity in theses analyses, and some evidence of horizontal pleiotropy or bias due to population stratification in the analysis among never smokers, but there is no clear evidence to suggest directional pleiotropy and the results are supported by the MR-Egger analyses.

***COPD***

The MR-IVW results indicate that increased NMR and smoking heaviness increase the risk of developing COPD among current smokers (OR = 1.37, 95% CI 1.20 to 1.56; OR = 13.41, 95% CI 7.53 to 23.88 respectively). However, the MVMR-IVW results indicate a protective effect of nicotine exposure when smoking heaviness is accounted for (NMR OR = 1.15, 95% CI 1.02 to 1.31) and a stronger effect of smoking heaviness when NMR was accounted for among current smokers (OR = 16.90 95% CI 6.82 to 41.84). These results are supported by the MR-Egger and MVMR-Egger results and there is no clear evidence of heterogeneity or directional pleiotropy or horizontal pleiotropy or bias due to population stratification among never smokers (where precise null effects are observed).

**References**

Buchwald, J., Chenoweth, M. J., Palviainen, T., Zhu, G., Benner, C., Gordon, S., . . . Tyndale, R. F. (2020). Genome-wide association meta-analysis of nicotine metabolism and cigarette consumption measures in smokers of European descent. *Mol Psychiatry*. doi:10.1038/s41380-020-0702-z

Bycroft, C., Freeman, C., Petkova, D., Band, G., Elliott, L. T., Sharp, K., . . . Marchini, J. (2018). The UK Biobank resource with deep phenotyping and genomic data. *Nature, 562*(7726), 203-209. doi:10.1038/s41586-018-0579-z

Evans, D. L. (1985). Cardiovascular adaptations to exercise and training. *Vet Clin North Am Equine Pract, 1*(3), 513-531. doi:10.1016/s0749-0739(17)30748-4

Howie, B., Marchini, J., & Stephens, M. (2011). Genotype imputation with thousands of genomes. *G3 (Bethesda), 1*(6), 457-470. doi:10.1534/g3.111.001198

Huang, J., Howie, B., McCarthy, S., Memari, Y., Walter, K., Min, J. L., . . . Consortium, U. K. (2015). Improved imputation of low-frequency and rare variants using the UK10K haplotype reference panel. *Nature Communications, 6*(1), 8111. doi:10.1038/ncomms9111

Kim, H.-G., Cheon, E.-J., Bai, D.-S., Lee, Y. H., & Koo, B.-H. (2018). Stress and Heart Rate Variability: A Meta-Analysis and Review of the Literature. *Psychiatry investigation, 15*(3), 235-245. doi:10.30773/pi.2017.08.17

Mitchell, R., Hemani, G., Dudding, T., Corbin, L., Harrison, S., & Paternoster, L. (2018). UK Biobank Genetic Data: MRC-IEU Quality Control, version 2 - Datasets - data.bris. *data.bris*. doi:doi:10.5523/bris.1ovaau5sxunp2cv8rcy88688v

O'Connell, J., Sharp, K., Shrine, N., Wain, L., Hall, I., Tobin, M., . . . Marchini, J. (2016). Haplotype estimation for biobank-scale data sets. *Nature Genetics, 48*(7), 817-820. doi:10.1038/ng.3583

Ware, J. J., Chen, X., Vink, J., Loukola, A., Minica, C., Pool, R., . . . Munafo, M. R. (2016). Genome-Wide Meta-Analysis of Cotinine Levels in Cigarette Smokers Identifies Locus at 4q13.2. *Sci Rep, 6*, 20092. doi:10.1038/srep20092

Yang, J., Lee, S. H., Goddard, M. E., & Visscher, P. M. (2011). GCTA: a tool for genome-wide complex trait analysis. *American journal of human genetics, 88*(1), 76-82. doi:10.1016/j.ajhg.2010.11.011
